# SARS-CoV-2 Vaccination Uptake in a Correctional Setting

**DOI:** 10.1101/2021.04.27.21252790

**Authors:** Justin Berk, Matthew Murphy, Philip A. Chan, Kimberly Kane, Jody Rich, Lauren Brinkley-Rubinstein

## Abstract

Between December 2020 and February 2021, the Rhode Island Department of Corrections offered SARS-CoV-2 vaccination to all correctional staff and sentenced individuals incarcerated in the state. During this initial campaign, 76.4% (1106 / 1447) of sentenced individuals and 68.4% (1008 / 1474) of correctional staff accepted and received the vaccine. This study demonstrates the feasibility and efficiency of vaccine implementation in a carceral setting.

## Introduction

The largest outbreaks of COVID-19 in the United States have occurred in correctional facilities.^1^ Correctional outbreaks have substantially contributed to community and statewide spread of infection.^2^ The rate of COVID-19 in correctional settings is five times that of the general population, and the age-adjusted mortality rate is nearly four times higher.^3^ Thus, vaccinating individuals who live and work in correctional facilities should be a high priority and is recommended by multiple organizations.^4,5^ Despite these recommendations, few states initially prioritized vaccination in correctional settings.^6^ Furthermore, vaccine uptake among correctional staff and incarcerated individuals is unknown.

The goal of the current study was to evaluate the rate of vaccine uptake among staff and incarcerated individuals at the Rhode Island Department of Corrections (RIDOC), the first statewide correctional system to offer vaccine to all sentenced individuals in February 2021.

## Methods

From the beginning of the pandemic, RIDOC collaborated closely with the Rhode Island Department of Health on addressing COVID-19 with aggressive testing and isolation procedures, mask wearing, surface sanitation, and ongoing education of staff and incarcerated individuals.

SARS-CoV-2 vaccines were initially offered starting on December 22, 2020 and first-dose administrations were completed on February 5, 2021. All second-dose vaccinations were completed by March 5, 2021. The RIDOC is a unified (combined prison and jail) statewide correctional facility that houses approximately 1,500 sentenced and 500 awaiting-trial individuals across six security facilities. The vaccine program focused on incarcerated people who had received a sentence after a criminal trial. This includes individuals typically housed in prisons. Staff (e.g., correctional officers) were concurrently vaccinated at the RIDOC through a parallel vaccine program. Among incarcerated people, a general system of “Rounds” prioritized vaccine allocation based on risk factors and/or security facility. All eligible individuals were offered vaccine on an opt-out basis. RIDOC nurses administered the vaccine. Two RIDOC public health nurses provided education on the vaccine and consent before the vaccine clinic day. Second doses were provided at appropriate time intervals.

In Phase 1, “highest-risk” individuals (age over 65 years old or over 55 years old with specific co-morbidities) were offered vaccine. In Phase 2, smaller facilities were offered vaccine in attempt to achieve herd immunity in those communities. Phase 3 included the largest remaining security facility: Medium Security. Phase 4 included all individuals who had previously tested positive for COVID-19 within 90 days and individuals who had initially declined but subsequently accepted. After completion of the 4 phases, vaccines continued to be offered by request and coordination with the local Department of Health.

Among corrections staff, individuals were vaccinated with an opt-in system, prioritizing high-risk correctional officers and individuals with direct contact with incarcerated people. Vaccine education was provided during morning “roll call” and an educational video was created by the Medical Director and made available on the intranet. Additional information regarding vaccine education and resources was sent via email to the entire RIDOC department.

## Results

During the six-week campaign, a total of 1,106 incarcerated individuals and 1,008 staff received the vaccine, 76.4% and 68.4% of the populations, respectively. (For comparison, the rate of influenza vaccination uptake at the RIDOC last year was 50.6%.) Table 1 describes the four phases of first-dose vaccination. Among Round 1 “highest-risk” recipients, approximately 9.1% declined the offer of vaccine. Among Round 2 individuals, rates of decline varied by security facility but were between 25% and 40% with an overall refusal rate of 35.6%. Round 3 included mostly individuals at the facility with the highest number of previous COVID-19 cases (Medium Security); the refusal rate was 17.1%. Round 4 rates of refusal were approximately 35%. Among staff, 1,008 employees or contractors received vaccine. A total of 466 of 1,474 individuals (31%) did not opt-in for the initial vaccine offering.

**Table 1.**
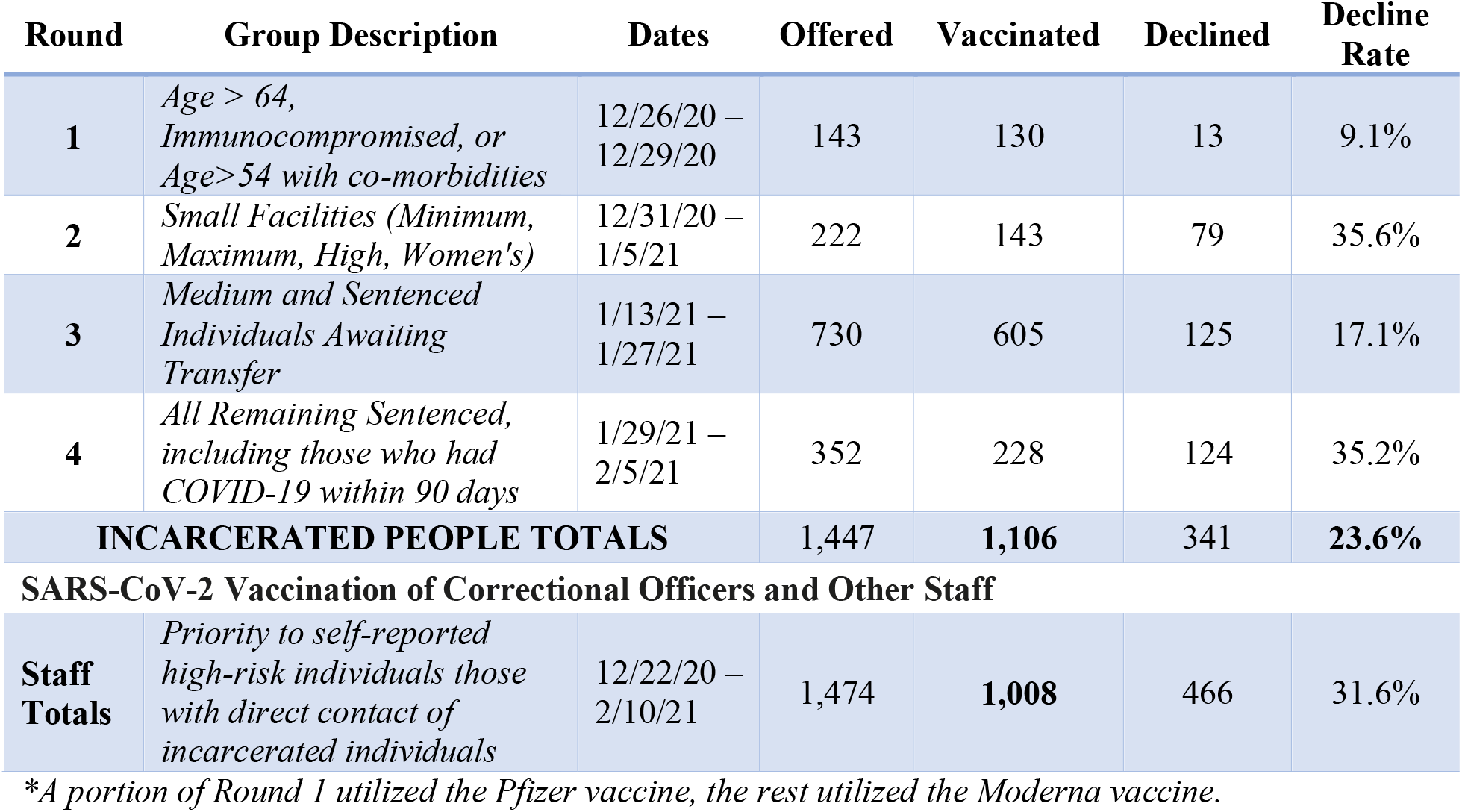
First-Dose SARS-CoV-2 Vaccination of Incarcerated People and Correctional Staff.

| Round | Group Description | Dates | Offered | Vaccinated | Declined | Decline Rate |
| --- | --- | --- | --- | --- | --- | --- |
| 1 | *Age > 64, Immunocompromised, or Age>54 with co-morbidities* | 12/26/20 – 12/29/20 | 143 | 130 | 13 | 9.1% |
| 2 | *Small Facilities (Minimum, Maximum, High, Women's)* | 12/31/20 – 1/5/21 | 222 | 143 | 79 | 35.6% |
| 3 | *Medium and Sentenced Individuals Awaiting Transfer* | 1/13/21 – 1/27/21 | 730 | 605 | 125 | 17.1% |
| 4 | *All Remaining Sentenced, including those who had COVID-19 within 90 days* | 1/29/21 – 2/5/21 | 352 | 228 | 124 | 35.2% |
| **INCARCERATED PEOPLE TOTALS** |  |  | 1,447 | **1,106** | 341 | **23.6%** |
| **SARS-CoV-2 Vaccination of Correctional Officers and Other Staff** |  |  |  |  |  |  |
| **Staff Totals** | *Priority to self-reported high-risk individuals those with direct contact of incarcerated individuals* | 12/22/20 – 2/10/21 | 1,474 | **1,008** | 466 | 31.6% |

\*A portion of Round 1 utilized the Pfizer vaccine, the rest utilized the Moderna vaccine.

**Table 2.**
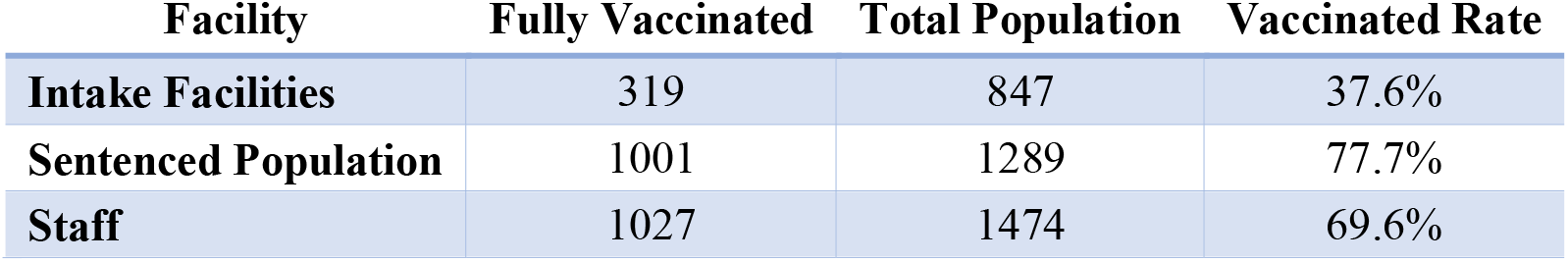
SARS-CoV-2 Vaccination of Incarcerated People and Correctional Staff (Four-Month Follow-up)

| Facility | Fully Vaccinated | Total Population | Vaccinated Rate |
| --- | --- | --- | --- |
| **Intake Facilities** | 319 | 847 | 37.6% |
| **Sentenced Population** | 1001 | 1289 | 77.7% |
| **Staff** | 1027 | 1474 | 69.6% |

Second-dose vaccines were administered approximately 28 days after the original dose. Due to logistics, some doses were delayed by 1 – 2 weeks due to vaccine delivery times and staffing availability. Three incarcerated individuals and six staff members who received their first dose of vaccine opted to not receive their second dose. During this time “over-pulls” and additional vaccine clinics were offered to incarcerated individuals and staff that ultimately did opt-in to receive vaccine on a rolling basis based on vaccine availability.

Four months after the first vaccine was offered on 12/22/20, 77.7% of the sentenced population was fully vaccinated and 69.6% of staff were fully vaccinated. At the time, 37.6% of the patient population at the Intake facility is vaccinated, though this is a transient population that is constantly changing. Future vaccination efforts for this population are planned to be single-dose vaccines. There were no significant vaccine adverse events.

## Discussion

Vaccination was efficient with a high acceptance rate of 70% - 75% among both staff and incarcerated people. This aligns with necessary immunization rates modeled to achieve herd immunity^7^ and is a departure from some concerns of high vaccine hesitancy rates including a recent CDC publication estimating a 45% willingness to receive vaccine among incarcerated people.^8^ Education and communication likely played an important role in mitigating refusals; efforts to increase vaccine uptake will continue. To our knowledge, the RIDOC is the first statewide correctional system to have offered SARS-CoV-2 vaccination to all sentenced individuals and staff.

The high acceptance rate in a correctional setting is particularly relevant given the increased risk of Covid-19-related transmission, disease, and death in this population.^3^ The pandemic has devastated correctional settings and the spread of disease in these facilities can catalyze transmission to their surrounding communities.^2^ Similarly, as Covid-19 has disproportionately affected communities of color, so too has rates of incarceration.^9^ Thus, by vaccinating incarcerated people, policymakers can target a high-risk and marginalized group, decrease community spread, improve equitable allocation to a marginalized group, and potentially reduce the health system costs of neighboring health systems. The successful vaccination of incarcerated individuals and staff in the state of Rhode Island demonstrates the feasibility and efficiency of widespread vaccine programming among those at high risk.

Vaccination of incarcerated people does have unique challenges. Rhode Island was able to coordinate the administration of second doses among the sentenced population without loss to follow up, but this was in part due to the small size of the state’s population. Additionally, the jail setting offers a greater challenge given the high turnover of the population often with individuals being released to the community before their second dose is due. The strategic implementation of single-dose vaccine may align with this unique environment going forward.

The success of this vaccination campaign exemplifies the importance of adherence to public health principles: vaccinate where spread and disease can best be prevented. Correctional settings should remain a priority in vaccination strategies during a pandemic and indeed offers an opportunity to target a high-risk, marginalized, and difficult-to-reach populations.

## Data Availability

The data is publicly available through media outlets or through requests to the Rhode Island Department of Corrections.

